# Fostering inclusivity: Navigating loneliness and wellbeing among Nigerian international students in the UK Higher Education

**DOI:** 10.1101/2024.06.10.24308728

**Authors:** Benjamin O. Ajibade, Petronella Mwalillanda

**Affiliations:** Northumbria University, Faculty of Health and Life Sciences. Department of Nursing, Midwifery and Health, Coach Lane Campus East, Benton. Newcastle Upon Tyne, NE7 7XA. United Kingdom; University of Sunderland, Faculty of Health Sciences and Wellbeing, 197 Marsh wall, London. E14 9SG. United Kingdom

**Author notes:** Correspondence author (BOA). These authors contributed equally to this work.

**Keywords:** Higher Education, Loneliness, International student Wellbeing

## Abstract

**Background:** Loneliness is a state of solitude or being alone. Research addressing loneliness has increased dramatically over the past two decades; however, despite the psychological state risks related to being lonely, the connection between loneliness and mental health disorders still needs to be sufficiently explored. Loneliness is when one’s relationships do not meet the psychosocial needs and expectations. Loneliness is a state of emotion that can be a barrier to social development and, at the same time, impact physical health and mental state. However, loneliness may be a distressing and pervasive experience, defined as the feeling that one’s desired quantity or quality of the social connection is unfulfilled.

**Method:** A qualitative primary research study was conducted for this study. The participants were selected using the purposive sampling method, and 8 participants of Nigerian origin were interviewed for this research. The participants interviewed were of Yoruba, Benin and Igbo origin; therefore, the results cannot represent the entire Nigerian population because it is missing the Hausa tribe. The data were analysed using the thematic analysis method. Ethical approval was obtained from the institution.

**Results:** The most salient themes were Voices of Loneliness: International Students’ Coping Strategies and Personal Journeys; Building Bridges: The Role of Community and Social Networks in Overcoming Loneliness; Exploring the Emotional Turmoil and Coping Mechanisms in a Foreign Landscape; Navigating New Realities: The Integration and Adaptation Challenges of International Students and Navigating Loneliness and Seeking Connection. The results showed that feelings of loneliness could result from emotional, psychological, or mental health and the problematic aspect of adaptation to the UK weather, as this greatly impacted their studies. Therefore, emphasis should be placed on understanding each student’s cultural background, assisting international students in examining their bicultural realities to recognise how clients internalise and adhere to culture considering their Nigerian cultural identities and values.

## Introduction

### Loneliness and Its Impact on International Students: A Comprehensive Overview

Loneliness, characterised by the feeling of unmet social needs in terms of quantity and quality of relationships, has gained increasing attention due to its profound impact on mental health and well-being. Researchers such as Hawkley and Cacioppo^1^ have defined loneliness as the distressing sensation arising from inadequate social interactions. Peplau and Perlman^2^ further elucidated this phenomenon as a disconnection between personal experiences and psychosocial desires. Diehl et al.^3^ emphasised the quantitative and qualitative loss of social interactions as a defining factor of loneliness.

Addressing loneliness has become a paramount concern for governments, as evidenced by the UK’s efforts to combat isolation.^4^ Initiatives like the ’Every mind matters loneliness campaign’ ^5^ underscore the importance of a collective approach involving individuals, communities, and professionals. Loneliness has been linked to adverse mental health outcomes, including depression, suicidal tendencies, sleep disturbances, and poor appetite,^6–8^, highlighting its potential to impact overall well-being.

Loneliness is particularly relevant in higher education, with a study by Neves and Brown ^9^ indicating that a quarter of university students reported feeling lonely most or all of the time.^10, 11^ The experiences of international students, especially those of non-EU origin, add a layer of complexity. The rise in their numbers, particularly from countries like China, India, and Nigeria,^12, 13^ necessitates attention to their unique challenges. The process of studying abroad often disconnects students from their familiar social support structures, contributing to feelings of isolation. ^14, 15^,

This research recognises the significance of addressing loneliness among international students, particularly those from Nigeria, as their transition into a foreign educational system can exacerbate feelings of isolation. Cultural differences, language barriers, and a lack of social support can compound the challenges faced by these students.^16^ The integration of cultural attachment theory into the transition process is crucial to ensuring a successful adjustment to the host country.^17^

The implications of loneliness extend beyond mental health, affecting physical fitness, academic performance, and overall satisfaction.^18, 19^ This research focuses on the experiences of Nigerian students in the UK, a group representing diverse cultural backgrounds and languages. ^20, 21^ Understanding their unique challenges is essential for developing targeted interventions to alleviate loneliness and enhance their academic and personal well-being.

By exploring the intersection of cultural adaptation, psychological well-being, and sociocultural adjustment, this study aims to contribute to a comprehensive understanding of loneliness among Nigerian students studying abroad. Through an innovative and inclusive methodology, this research seeks to uncover strategies that can effectively address loneliness and promote a supportive and enriching university experience for international students, ultimately fostering an inclusive and diverse academic community.

## Methods

The choice of a qualitative research methodology was made for this study as it delves into how individuals perceive themselves and their surroundings, and how they attribute meaning to their daily experiences. ^22, 23^ Qualitative researchers often adopt an interpretive or critical social science approach, using logical methods and following non-linear research paths. They prioritise in-depth examination of cases that naturally emerge in social contexts.^24, 25^

### Aim and Objectives

This study aims to explore the impact of loneliness on Nigerian students studying in the UK. The study objectives are as follows:

- Investigate Nigerian students’ perspectives on loneliness and their coping strategies.
- Critically assess the bio-psycho-social implications of loneliness on Nigerian students.
- Examine the effect of loneliness on the psychosocial well-being of Nigerian international students.
- Compare students’ perceptions of loneliness between their experiences in a foreign country and Nigeria.
- Evaluate how Nigerian students perceive and overcome loneliness, contributing to knowledge co-construction.

### Sampling Method

A purposive sampling method was employed.^26^ This non-probability approach involves selecting individuals from the target population based on practical criteria such as accessibility, proximity, availability, or willingness to participate.^27^

### Purposive Sampling

Purposive sampling emphasises reaching saturation, which means continuing sampling until new meaningful information ceases to emerge. Convenience sampling is cost-effective and easily accessible, often involving readily available subjects.^28^ The underlying assumption is that the target population shares homogeneity. While various sampling methods may yield similar results, purposive sampling aligns with this research study.

### Data Collection Methods

The primary objective of this qualitative research was to gain profound insights into students’ experiences and the effects of loneliness.^29^ Originally planned as face-to-face interviews, the COVID-19 pandemic necessitated a switch to online interviews, adhering to government guidelines. Participants were informed of this change, and ethical approval was secured from the university before data collection commenced. Data collection is a critical research phase, as the gathered data contributes to the theoretical framework’s understanding. The choice of data collection method is pivotal, and purposive sampling, also known as judgment sampling, involves selecting participants based on specific qualities they possess.^30^ While some prefer diaries for data collection,^31, 32^ face-to-face interviews are favoured by others due to their capacity to yield extensive open-ended responses.^28^

### Sample Size

Eight participants were interviewed for this study. Crouch and McKenzie^33^ suggest that a qualitative study with fewer than 20 participants allows researchers to establish and maintain close relationships, facilitating open and honest information exchange. This approach helps mitigate biases and threats to validity inherent in qualitative research.

### Ethics statement

To ensure the integrity and ethical standards of the study, ethical approval was sought from the university ethical committee in compliance with the university ethics policy. The process involved submitting a detailed research proposal outlining the study’s objectives, methodology, potential risks, and benefits to participants, including the strategies for ensuring participant confidentiality and data protection. The participant information sheet was given to participants before requesting their consent to participate in the research. Informed consent was obtained from all participants before their involvement in the study. Consent was documented through a signed consent form, ensuring that participants fully understood and voluntarily agreed to participate in the research. The research participants’ autonomy was observed and were informed of their freedom to withdraw at any time without penalty, measures were taken to protect their privacy and confidentiality.^67, 68^ Confidentiality was maintained by anonymising the participants with pseudonyms, and the data was kept secure and encrypted in compliance with Data Protection Act.^69^ By adhering to these ethical standards, the research upheld the principles of respect, beneficence, and justice, thereby safeguarding the rights and well-being of all participants.

## Results

Data analysis involves a comprehensive process of reading, re-reading, restructuring, examining, and presenting data, with the potential for seeking further insights from participants to enhance clarity and depth.^34^ This approach aims to uncover the significance of the gathered data.^35^ The data will be transcribed following the step-by-step thematic Analysis method, facilitated by the use of the QUIRKOS software.^36^ Thematic analysis serves as a valuable tool for dissecting and understanding data, commonly employed across various qualitative research designs. It aids in identifying, interpreting, and evaluating patterns or themes within the collected information. While Caulifield^37^ presents diverse approaches to conducting thematic analysis, Braun and Clarke’s six-step framework is widely adopted. This process encompasses becoming familiar with the data, coding, theme generation, theme review, theme naming and definition, and finally, compiling a comprehensive report.^35^

Among the identified themes, the most prominent are Voices of Loneliness: International Students’ Coping Strategies and Personal Journeys, Building Bridges: The Role of Community and Social Networks in Overcoming Loneliness, Exploring the Emotional Turmoil and Coping Mechanisms in a Foreign Landscape, Navigating New Realities: The Integration and Adaptation Challenges of International Students and Navigating Loneliness and Seeking Connection.

### THEME 1 – Voices of Loneliness: International Students’ Coping Strategies and Personal Journeys

The data provide perceptions of the diverse experiences and emotional impact of loneliness among international students. The participants jointly illustrate the multidimensional nature of loneliness, thereby showcasing a range of coping mechanisms, emotional experiences, and perceptions of social isolation.

### Navigating Solitude and Intellectual Pursuits

One of the participants expressed that when they feel lonely, they tend to retreat to their room and engage in intellectual activities like reading educational materials and novels. They don’t outwardly express their emotional loneliness but rather channel it into a productive pursuit. This suggests that they find solacement in intellectual endeavours when dealing with loneliness.

> *“…I didn’t know where to go, so sometimes I ended up being in my room eerm. That is how I expressed my loneliness but the emotional aspect of it; I don’t personally express it physically. I kept reading educational journals and novels …” (Gabriel)*.

Another participant highlights the loneliness experienced during break times when there is no one to socialise with. Their quote features how brief periods of aloneness, such as break times, can contribute to feelings of loneliness, stressing the importance of social interactions in mitigating such emotions.

> *“…*and *if there is no one to hang out with at break time, then you are by yourself…” (Stephanie)*.

Some other participants discussed shutting down emotionally due to their inability to connect with a specific group, causing self-blame. This internal struggle with loneliness significantly impacted his coursework and overall well-being. His quote demonstrates the complexities of loneliness, including its association with self-blame and its tangible effects on academic performance.

> *“… but I completely shut down because I could not see myself engaging with people from that group. [pause] I kept blaming myself …… errm most of the time …. which was unfair on myself because I didn’t know … and it made me feel lonely if I may say … er because it impacted on my course work (William)*.

Some participants shared the impact of loneliness on their concentration and academic performance being in a foreign country, emphasising how thinking about family and friends affected their ability to focus. They expressed feelings of isolation and apprehension, particularly regarding their surroundings and cultural differences. However, they managed to cope with these feelings by reaching out to their family when lonely.

> *“..It affected me then because I was unable to concentrate on my studies because whilst I was thinking about my friends, families or their facts, it really affected my academic performance then, I wouldn’t lie, but I managed to survive by probably as in contacting them when I felt lonely..” (Mo)*.

> *“So I feel oh., why did I come to [UK], so I feel so lonely about it. How things are going on around me in a foreign land” (Brenda)*.

Other participants expressed loneliness as the absence of a social network, involving family and friends. They described it as a state of isolation where individuals are unable to engage with others. This sheds more light on the social aspect of loneliness.

> *“..Loneliness is when someone has not got erm social network [pause]. This can be in terms of family and friends around the person, and the person is more or less experiencing errm… isolation of some sort, not able to engage er..” (Gabriel)*.

## The Impact of Weather and Climate on International Students’ Adjustment

It was underscored that people often receive warnings about the UK’s weather, but it’s challenging to fully prepare for it. The newcomers are caught off-guard due to this unpredictability, leading to difficulties in adjusting to the climate.

> *”..when you are about to come to the UK, people would tell you all sorts of things UK is this UK is, that but you cannot fully prepare I tell you” (Princess)*.

Some participants reflect on the encounters faced by international students regarding clothing choices. It was remarked that bringing inappropriate attire for the UK’s colder climate can lead to discomfort and a reluctance to go outside, hypothetically contributing to feelings of seclusion.

> *“that contributes a lot to loneliness; you can’t just wear just an ordinary T-Shirt and go out. It affected me a lot as a student… because I brought summer clothes and then wow [giggles] I could not believe it …. It was very cold. I was going to Uni, and sometimes I was even feeling like just staying in bed.” (Mo)*.

Some of the participants indicated the necessity of wearing multiple layers of clothing in the UK, which differs from their home countries where lighter clothing may suffice. This cultural difference in dressing can be a significant adjustment for international students.

> *“..In winter, back home, we wear a sweater or a cardigan, but here we wear a cardigan, we wear a top, you wear trousers, you wear tights, you wear a coat which is for me double layer upon layer. Back home, we don’t have something like that, so it is easy there, but here [giggles]” (Brenda)*.

A participant recognises the effect of weather on mood, with darker and colder days potentially reducing the motivation to go out and socialise. This weather-related mood change can contribute to feelings of loneliness.

> *“I know for a fact that weather can affect mood; I think it affected me. [chuckled] If it’s dark outside, you have less pleasure to go out than when it’s bright. First, I found the dressing very horrible, layer upon layer of clothes” (Stephanie)*

Another participant emphasises the significant geographical and environmental differences between the UK and international students’ home countries. These differences, including the harsh weather and unfamiliar terrain, can create additional challenges for adaptation.

> “[Prolonged loud laughter] ……… geographical background you know …definitely. It is really different….. You have to face the challenges of weather, having to wear lots of clothes to keep you warm … and err … It’s not a place of a familiar terrain, but its quite a difference” (Amy).

Some of the participant accentuates the feeling of coming to a place that is entirely different from what one has experienced before. They reinstated that the initial shock of experiencing a harsh winter and unfamiliar conditions can make the adjustment period particularly difficult.

> *“.. [long Pause] Right …. Ok… this is something like you are coming to a place that you have never experienced or known, so witnessing it first, it’s obviously winter being the harsh weather, very cold and not used to that erm it was difficult..” (Gabriel)*.

### Understanding the Nuance of Loneliness Among International Students

The participants emphasised that loneliness among international students goes beyond physical isolation, underscoring the significance of meaningful social connections and emotional support. They stressed that the presence of individuals alone is insufficient, and having a supportive network and a sense of belonging is vital for overcoming loneliness. Trustworthy relationships and a strong community or support system are crucial in preventing loneliness among international students.

> “you can have people around you and still be lonely,” (Princess)

> *“loneliness, I would say, when one feels … erm …. Maybe you have’t got friends or people you can rely on when you feel isolated” (Amy)*.

> *“Ok .. Er … My understanding of loneliness is when you have no network support, or there is nobody there in your network” (William)*.

Another student’s experience of loneliness in their living situation, particularly when left alone in their room, points to how living arrangements can impact feelings of loneliness. It shows that the physical environment, such as living in a small, isolated space, can exacerbate feelings of loneliness, which in turn can affect academic performance.

> “*….I experienced a bit of loneliness because I have my own little room in which I share a bathroom with my colleagues in a flat. then [phew] when everybody just goes out, I feel lonely in my little room........which sometimes affects my education or academics...” (Mo)*.

### Cultural Differences and Their Impact on Loneliness in International Students

Participants delve into the impact of cultural backgrounds on loneliness among international students, emphasising the challenges of adapting to a new cultural environment. They note that difficulties in understanding and adjusting to a different culture can contribute to feelings of isolation, particularly when cultural beliefs diverge significantly from those in the new environment, fostering a sense of disconnection.

> *“..so in that aspect, your cultural beliefs could more or less impact or make you feel* lonely *in a new culture or a new environment because you find that its hard to actually understand that the culture you are in”… (Gabriel)*

Nigerian ethnic background illustrates how moving between different cultural environments, even within one’s own country, can lead to feelings of isolation. This is aggravated when moving to an entirely different country like the UK, where even though there might be a newfound sense of unity among people from the same home country, the initial cultural differences can still contribute to a sense of loneliness.

> *“Each Ethnic background has got a college or Uni, so if you tend to move from an Igbo area to the Yoruba area, you feel isolated because first, you need to learn the language although there is a common language we use .* …..*erm.. But they would know through your accent that you are not part of them so you will be isolated definitely. Once you leave the country [Nigeria], the attitude towards each other changes once you leave Nigeria and come to the UK, if you see someone from Nigeria no matter where the person comes from, Igbo, Yoruba” (Mo)*

Cultural norms, particularly around marital roles and expectations can contribute to loneliness. In this context, cultural expectations around submissiveness and not discussing domestic issues can lead to isolation and a lack of support, particularly for women.

> *“… for instance, you know, married couples, things are not going well, when they have their differences, and they have their domestic issues .. So the husband stops talking to the wife, the wife is not able back home in Africa, you know women are expected to be submissive to their husbands and not to report their husbands to anybody” (William)*

### Language and Accents: Navigating Communication Barriers as an International Student

International students encounter challenges such as language barriers, accents, and variations in educational systems, impacting their adaptation and contributing to loneliness. The struggle is not solely with language but also with local accents, hindering communication, and causing misunderstandings, frustration, and isolation. This extends beyond language proficiency to unfamiliar local dialects, slang, and idioms, making full integration into social circles challenging.

> *“ for me … it was the language barrier… or …[pause] …. Can I rephrase that please, it’s the accent I did not understand” (Stephanie)*.

> *“well, not a massive impact … but a little bit, but again it is because you are coming from the country where you don’t speak their language even though you speak English in Nigeria, but you don’t speak in a way” (William)*.

There is a difference in the educational approach to English in Nigeria compared to the UK. The implication here is that even though English might be taught as a primary language, the level of exposure and the context in which it is used can vary greatly, affecting students’ confidence and ability to communicate effectively in a new educational environment.

> *“Back home is different from the way we study here. Our English might be the first language, but it’s never the first language in Nigeria, or they make everyone [sigh] study English, let me put it that way.” (Mo)*.

It’s difficult to make new friends and engage in social activities due to language and cultural differences. This barrier is not only about communicating in English but also about understanding cultural cues and norms that are essential for social integration. The feeling of loneliness is worsened by the lack of physical presence of friends and family from home, with digital communication being the only link to their support network.

> *“.. So [pause] it was more difficult to settle in and make new friends who were predominantly white people’s background; you rarely see people of the same ethnic origin. So it was difficult. I was more or less going to school and coming back … or my main communication with friends and family was just through the phone, but physically it was difficulty to engage and eerr in social activities so I would say that this is my first experience to feeling lonely since arriving in the UK,..” (Gabriel)*.

### THEME 2: Building Bridges: The Role of Community and Social Networks in Overcoming Loneliness

The theme reiterates the importance of social connections, community engagement, and the pursuit of common interests in mitigating feelings of loneliness, particularly among international students. It reflects on various aspects of forming relationships, the challenges, and solutions to feeling isolated, and the significance of finding a sense of belonging in a new environment.

### The Power of Workplace Friendships

The friendships formed in the workplace can significantly enhance job satisfaction and create a welcoming and enjoyable work environment, blurring the lines between professional and social spaces. There are individual differences in how people approach friendship and humour, and openness are important in forming new connections.

> *“I made friends with girls at work; I enjoyed my job, I loved what I was doing and coming to work was as if I was coming to see my friends, and we were working together ” (Princess)*.

> *“I tend to make friends; I am a very jovial person. It is a difficulty for some people to make friends. I tend to make friends easily. I tend to crack a lot of jokes, so making friends is easy” (Mo)*.

### Unity in Diversity

This reflects on the unique situation of international students from countries like Nigeria, finding unity and a sense of community with fellow nationals, overcoming ethnic and linguistic differences when abroad.

> “*Once you leave the country [Nigeria], the attitude towards each other changes, once you leave Nigeria to come to UK if you see someone from Nigeria no matter where the person comes from Igbo, Yoruba etc. As long as the title of the nationality is Nigeria, you are one, and you tend to click together. You leave your differences back home” (Mo)*.

> *“Any activity that would bring people of the same culture or ethnic together is not always possible, but if you spend some time and research like things that are happening that might be of interest for you in terms of social gathering or social activities, yeah, obviously there could be more that could be done but when you are in a place like London, it’s a bit easier because you can meet other people.” (Gabriel)*.

### Cultural and Social Gatherings

It is important to seek out and participate in cultural and social activities that can connect individuals with similar backgrounds and interests, emphasising the role of effort and research in finding these communities, particularly in multicultural cities like London.

> *“But subsequently things were much better because once you start to develop that social network and meeting other people, you tend to find eeerr common social activities that you enjoy by meeting other people with the same ethnic origin” (Gabriel)*.

### The Importance of Local Networks

The emphasis on registering with local networks and communities showcases the necessity of having a support system in place for international students to prevent loneliness and offer a sense of belonging.

> *“I think it’s important to have students registered on their local network because it is important…. [pause] You can’t feel lonely … You might think, oh! I can do it by myself, but when the chips are down, you realise that, oh ghosh …. This is bigger. Say, for instance, being in the Nigerian community in every part of London, say, for instance, a student comes to study in East London” (William)*.

### Shared Interests

One of the participants revealed knitting as a communal activity back home which points to the significance of shared interests in forming connections and the potential for universities and communities to facilitate interest-based groups for students.

> *“Maybe they should ask students their interests, especially where they came from; for me, I liked knitting with other people when I was back home during my spare time” (Stephanie)*.

### Challenges and Solutions to Loneliness

The narrative transitions from discussing the challenges of loneliness and the vulnerability it can create to the transformative power of developing social networks and finding common social activities.

> *“I think… others might indulge in doing [pause] all sorts of activities [chuckles], and that can lead them to be vulnerable towards others just because they are lonely. Things like online dating” (Princess)*.

> *“Loneliness can affect students with their mental state, which can have an impact on their learning. Because if you have no one to talk to, no family to talk to say to you that you know that sometimes regarding that, what people tend to, especially Christmas, for example, people tend to go to their family members, get together and have a meal” (Mo)*.

### Communal Living and Cultural Perspectives on Loneliness

There are cultural differences in perceptions of loneliness, with a particular focus on the communal living arrangements typical in Nigerian society, which contrasts with the individualistic lifestyle more common in the UK.

> *“Loneliness is really not an issue back home. errm …. [pause] Nigerians believe in communal living” (Amy)*

> *“You have a family of 3 generations living together, so you are living with your cousins, and people in school didn’t know they were my cousins and people in school did not know they were my cousins because we were like sisters.”(Princess)*

> *“Loneliness can affect students with their mental state, which can have an impact on their learning. Because if you have no one to talk to, no family to talk to say to you that you know that sometimes regarding that, what people tend to, especially Christmas, for example, people tend to go to their family members, get together and have a meal” (Mo)*.

### Religious and Ethnic Community Support

The role of religious communities and ethnic associations in providing a welcoming space for new arrivals, facilitating social integration, and offering emotional and spiritual support is a recurring theme, with personal anecdotes from Princess, Amy, William, and Gabriel illustrating how these spaces can serve as vital social and support networks.

> “*What people tend to do is because the majority of our people are Christians. So when they move to a new place, they locate the Pentecostal church in that area. If you go to church on Sundays” (Princess)*.

> *“I think in terms of the students themselves, I think if they can try to identify or if it is easy for them to identify places where they feel they belong… people use church activities as well to socialise or which is helpful” (Gabriel)*.

> *“Sometime going to different places for church groups …Because in my community I used to go to a Pentecostal church and religion is respected so much … I felt part of the community.” (Amy)*.

> *“I believe churches do the same as religious organisations; yeah, they could find a place. Go to church [pause] and meet people of err of the same background.…so that they are not completely left out ..So students should endeavour to do that” (William)*

### Theme 3: Exploring the Emotional Turmoil and Coping Mechanisms in a Foreign Landscape

The data paints a poignant picture of the emotional challenges faced by international students, especially during the winter months. The feelings of withdrawal, moodiness, and the visible impact on mental well-being are intricately described by the participants.

### Impact of Winter Darkness

The mental toll of winter darkness explains how the early onset of darkness can create a stark realisation of being in a new place. The physical and emotional toll, with extreme cold leads to a sense of numbness and even tears that go unnoticed due to the moist surroundings. The demotivation caused by dark and cloudy days affects visibility and the willingness to go outside.

> *During winter, erm… it becomes dark, say from 3:00pm, 3:30pm. About that time, it gets dark, so it is … You just know that you are in a new place. You know …Things are really different. It affects you mentally, at times, you even lose track of time. And erm” (Amy)*.

> *“I was frozen, you know, like you don’t feel any part of your body [pause] err the blood drained away from the extremes, eeerr it was extremely difficult ….. I cried …..I went to the bathroom and cried on my way back home,I cried In fact, when you cry, people don’t even know that you are crying because everything looks moist” (William)*.

> *“I wouldn’t lie to you when the weather is dark all the day when you wake up with no one to talk to and it’s even dark, very cloudy outside, low eeerr, visibility is abysmal, you don’t have to motivation to go out, you might know .. Especially if you have no school or no” (Mo)*.

### Loneliness and Emotional Struggles

The data postulated a sense of sadness and loneliness for international students, feeling that nobody wants them and lacking support. The emotional impact of loneliness describes the need to make unnecessary calls during lockdown to combat feelings of isolation. On the other hand, a participant reveals the emotional toll of being away from family, with moodiness setting in if they don’t connect with them regularly.

> *“I feel [pause] sad sometimes as an international student that I came over here. So, then I know gradually I am going to be ok. I feel bad for being here, yeah; I think like nobody wants me …maybe that’s what I know …. That loneliness is all about how I feel that things are going on around me … nobody is there to help me. That’s the way I see loneliness to be” (Brenda)*.

> *“then I started making unnecessary calls to see whom I could talk to … like when we were in lockdown during the COVID period, I. er .. I call my friend, and sometimes I would realise that oh my God, it’s 9 on’clock in the morning, but because I have been up since 6, I would just pick up my phone and ask where are you”(Princess)*.

> *“But before, I used to call home almost every day and speak with my family. If I don’t call [pause] … I would become very moody and start worrying in case maybe something has happened…Er.. If they answer and we will all be happy, and they would tease” (William)*.

### Communication and Coping Mechanisms

The importance of communication emerges as a recurring theme. The participants reflect on the significance of reaching out to people back home, even if they may not fully understand the struggles abroad. They also mention making calls during lockdown as a coping mechanism. Others acknowledges the potential negative impact of relying on addictive substances, such as smoking and alcohol, as a way to cope with isolation and stress.

> *“But before, I used to call home almost every day and speak with my family. If I don’t call [pause] … I would become very moody and start worrying in case maybe something has happened…Er.. If they answer and we will all be happy, and they would tease” (William)*.

> *“So the only person you can talk to are the people back home, and maybe they have no experience of what you are going through in Europe…so you will continue feeling lonely and especially when you have course work to submit you don’t have much friends, and you don’t know whom to trust” (Brenda)*.

> *“then I started making unnecessary calls to see whom I could talk to … like when we were in lockdown during the COVID period, I. er .. I call my friend, and sometimes I would realise that oh my God, it’s 9 o’clock in the morning, but because I have been up since 6, I would just pick up my phone and ask where are you”(Princess)*.

### Discrimination and Seeking Support

The participants have various views on discrimination ranging from personal experiences of discrimination, showcasing the diversity of perspectives to the importance of letting people know about mental health struggles and advocating for timely intervention to prevent deterioration. Although there is positive experience with the University Wellbeing team which underscores the significance of institutional support. One of the participants discussed the long-term consequences on mental health, highlighting how the initial difficulty in sharing experiences and forming connections can escalate into stress, impacting mental health over time.

> *“I am actually ok. You know this doesn’t mean I am not ok and the more that carries on the more it becomes a major problem” (William)*.

> *“And again, people are different again. but.. I didn’t .. to be honest. I don’t think I was subject to any discrimination” (Mark)*.

> *“To be honest, I was introduced to the University Wellbeing team; they told me if I had any problems like mental health issues or loneliness or financial issues that I should let them know, and they will give me support, and I was having financial problems and like I told you I was having loneliness and I think I was having a little bit of mental health problems” (Brenda)*.

> *“…some find it hard and err… It then starts to affect them with their mental health not having someone to share may be their good times and their bad times and their difficulties, and it then builds up to stress and affects their mental health ”(Gabriel)*.

### Theme 4: Navigating New Realities: The Integration and Adaptation Challenges of International Students

The theme collectively narrates the multifaceted challenges faced by international students as they navigate academic environments, social integration, and cultural adjustments in a new country. These extracts shed light on the initial struggles with academic tools, the importance of social support systems, and the process of cultural adaptation, each contributing to the broader experience of studying abroad. The narratives collectively underline the difficulties of adjusting to a new academic system, the critical role of social and cultural integration efforts by universities, and the personal journeys of cultural adaptation. They highlight a shared need among international students for comprehensive support systems that address academic, social, and cultural challenges, facilitating a smoother transition and enriching their educational experience abroad.

### Academic Adaptation and IT Skills

There are gaps in participants IT skills. This is evidence during introduction to Turnitin which highlights the gap in digital literacy and academic practices faced by students from different educational backgrounds. The students experience emphasises the transition from paper-based work to digital submissions, accentuating the need for targeted IT training. A participants narrative about their handwritten essay being rejected illustrates the stark differences in academic expectations and the necessity of acquiring new skills, such as typing.

> *“say turnitin… [pause] this is new to me as I have never heard of that [laugh] pause… but it is what it is ….ok in terms of the IT skills I talked about earlier if there would be a kind of a training, so IT supports immediately eerr OK after induction” (Amy)*.

> *“..But again [pause] not knowing how to use Turnitin assignment work and not able to operate IT skills was a tough …. You see [pause] In Nigeria we are used to writing on paper in a class, but when I came here, most of the work was computer-based…. I lacked some skills, and I felt I was not helping myself …” (William)*

> *“and I remember writing an essay at the time handwritten, and it was dejected. [laugh] And I had to go and learn how to type” (Mark)*

### Social Integration and Cultural Celebrations

Some participants suggest the organisation of social gatherings and cultural evenings as a means for students to connect with peers from similar backgrounds, thereby fostering a sense of community. This expresses the broader goal of celebrating diverse cultures without losing one’s identity, and stresses the importance of cultural understanding for smoother integration.

> *“hold social gatherings for students to mingle and find people they will relate to …so if they can provide that for me, then I will feel close to home… maybe say… a student that comes from… err what example can I give… India” (Stephanie)*.

> *“ I don’t know if they have it, I‘ve not seen it that they can say; people from Nigeria community. So they will have a social evening maybe once a month where all Nigerians will come together, or all Chinese students will come together once a month, so that would be helpful.” (Princess)*.

> “*So it’s all about celebrating other people’s culture and traditions, you know, engaging with whatever is out there, not losing your personality” (William)*.

> *“I think the university should recognise that people come from different backgrounds. If you go anywhere, you need to understand the culture, because that will help with integration and ask questions” (Mark)*

### Induction and Continuous Support

The participants discussed the potential of a well-structured induction process to alleviate initial stressors, emphasising early support in academic and practical matters. They further proposes that universities should monitor students’ academic performances closely, providing timely interventions to address any issues. Navigating the university environment, from accessing resources to understanding the local job market, highlight the need for ongoing guidance beyond the academic territory.

> *“the induction process, it would really definitely help to alleviate the stress some of these students experience… you know…. having to deal with schoolwork.” (Amy)*.

> *“your first …. What is it called … Your first performance will show, and they will know the type of grade that you will be getting when they see that the grade is going down, they should be able to put something in place; they should determine and talk to that person what is going on” (Mo)*

> *“..and sometimes the environment itself, not knowing how to access things within the university and other things like email, SKILLS ……. Moreover, [pause]… accessing temporary jobs, what to do, what not to do [chuckles]” (Stephanie)*

### Cultural Adaptation and Navigational Challenges

Participant’s reference to bringing familiar seasoning items like Maggie cubes (seasoning/spices) and dry fish illustrates the tangible aspects of cultural adaptation related to food preferences. While some of them have humorous reactions to local cuisine by indicating they are going to eat a leaf, others decided to source local food instead of relying on shipments from home capturing the initial culinary adjustments and the eventual acceptance or adaptation to new food cultures.

> *“Before, if they tell me that somebody is coming here, then I would write a list of what they should bring, especially Maggie cubes for cooking and dry fish and dry fish” (Stephanie*)

> “I was shocked and asked myself am I going to eat a leaf?” (William)

> *“So that made it a bit quicker. I like my local food …[pause] I was alone then so send food for me to me .. I look at it as a lot of stress … so I can I get it here*

> *.. Then why not?” (Mark)*

### Theme 5: Navigating Loneliness and Seeking Connection Emotional Burdens and the Need for Support

Participant’s quotes highlight the emotional strain of loneliness and the deep-seated need for companionship and support. A participant feels the absence of someone to confide in during troubling times, while others describe the despair and lack of happiness that can lead to an existential crisis. This demonstrates the critical need for a supportive social network that can offer help and understanding.

> *“especially when I have something that is bothering me… there is nobody around me to … for me to tell them that this is my problem can you help me …. so sometimes I will have that feeling inside me to have somebody around me to talk to” (Brenda)*.

> *“like feeling down… dejected, eeer ……. [Pause] having lack of happiness in their lives …. having nothing that makes you want to live another day” (Stephanie)*.

### The Power of Communication and Connection

Open communication is important in alleviating loneliness. It was pointed out that without communicating one’s struggles, others remain unaware and unable to offer help, potentially leading to worsened mental health. There is an emphasis on maintaining regular contact with family, using long chats and calls as vital strategies to combat loneliness and stay connected.

> *“let people know, and the intervention that is appropriate can be prefer because if you don’t talk …[pause] …people don’t know what is going on with you. ……..and then the more you kept quite, the more things get to deteriorate to a point that you can become depressed” (William)*.

> *“ If one can make friends, it will also go a long way and err… try to find out yourself things that will be of use…. You get involved in some activities that will take off that feeling of the burden of loneliness err I do call my family back home on a regular basis because I need to know what is going on over there. At the same time, I am also overcoming my loneliness … [giggles]” (Amy)*.

> *“for me, I use the time to spend with my family… I just call them sometimes, and we have these long chats [pause] other times when I don’t feel like talking to anyone because I also feel that sometimes, then I will do a lot of readings” (Mo)*

### Distractions and Engaging in Activities

Distractions and activities were used at various times to cope with loneliness. While some engage in watching films and immersing themselves in various hobbies to distract from loneliness, others engage in solitary but fulfilling activities like reading, which help them manage feelings of isolation even if they don’t completely eradicate them.

> *“…for me [pause], I feel lonely if I am by myself even for a few hours… then I start to feel lonely. but …. But [pause] I distract myself with all sorts of things” (Stephanie)*

> “And I watch movies, I like to err.. a lot of love stories and you know those with the black heritage, like Black American films” (Princess).

> *“I have experienced loneliness; it wasn’t severe. I have tried to get myself busy with other activities even though I still feel lonely because I am not in touch or likely to be in touch with other people.” (Gabriel)*.

### Visible Signs and Social Withdrawal

The physical manifestations of loneliness are poignantly described by some participants. Some participants reveal that withdrawal and silence signal emotional state to those familiar with them, indicating the importance of recognising such signs in addressing mental health. Loneliness was broadly defined as a state of isolation that involves a lack of all forms of social contact, underlining the severe impact of loneliness on individuals’ lives.

> *“.So if I can’t hear anybody, then I will go and close myself somewhere; sometimes I go to my bed, or I become so quiet even when people come around me* ….*I will not even watch the television, nothing [pause], trust me. That’s how I express my loneliness, and if anybody that knows me very well comes close to me, that time you will know that there is something wrong with me; somebody that is always chatting and laughing with me, especially my brother, so when you see me am quite like that you know that something is wrong..” (Brenda)*.

## Discussion

This research aims to explore the impact of loneliness on Nigerian students while studying in the UK. International students constitute an increasingly relevant and vital source of diversity on college campuses. Through their home culture and multicultural experiences,^20^ they enrich the cultural diversity of universities. ^38, 39^ The changes to the routines, diets, social environment, geographical environment, and perceived demands of new students can cause extreme homesickness.^40^ Although some international students have prior experience travelling away from home without guardians, all students must face the difficulties of handling their lives individually because the result of dysfunctional communication, stress, anxiety and depression is loneliness. ^41^

According to the literature, Diehl^3^ explains that there are multiple transition periods in life and that they may be critical.^3^ Ajibade and Hayes^17^ opined that transition is individualised and takes different times to transition to a new environment. Hence, some participants queried their decision to study in a foreign land. Another scholar explains that mental health issues are highly prevalent in university student communities, particularly among those transitioning from secondary to tertiary education.^42, 43^

According to another report, a lack of competence in a host country’s language has increased anxiety for international students.^44^ As illustrated in the literature, three fundamental causes that may contribute to the cause of loneliness are a sense of belonging to a new community or team, feeling alone, and self-denial.^45^

One of the factors reported by some students was preconceptions about the weather in the UK. One respondent described how she only brought summer clothes and felt helpless and withdrawn. This demonstrates why in winter times, there are high rates of suicidal ideation and heightened decline in mental health.^46^ About the comprehensive literature obtained, some studies have identified weather as one of the critical problems for some students. For example, research carried out in Malaysia described climate-related issues as one of the most challenging factors faced by international students. ^47^

International students expressed a language barrier as an additional concern about studying in a foreign country. The participants acknowledged that although English was the official language in Nigeria, ^21^ the accent was completely different in the UK, which is compounded by the various accents in the UK, thereby causing more confusion for Nigerian students. This resulted in the participant being mocked due to their accent. Research from other sources concluded that the obstacles and challenges faced by international students are at an all-time high.^45^ Most students expressed isolation and lack of contact as major contributing factors to loneliness. The COVID-19 pandemic was seen as a significant precursor to increased social isolation and loneliness due to the lockdown in the UK. Hunley^48^ explains that students may also feel detached from the community around them.

Loneliness has been described as unpleasant feelings where there is a difference between what one desire and what one has in terms of emotional affection and intimacy. This study highlighted transitioning issues^49^ as the main theme, with sub-themes helplessness, homesickness, acceptance, and psychological and mental health issues as some of what the participants expressed. According to Diehl et al.,^3^ there are two distinct forms of loneliness: a lack of close and personal relationships that contribute to relational loneliness and a lack of a social partner network that leads to social loneliness. For example, psychological isolation emerges after a partner’s divorce or death. At the same time, social loneliness exists when someone is not socially integrated, such as in a group of friends with mutual interests. International students are vulnerable to loneliness because both types of ties are missing^50^

Students can experience psychological stress while studying abroad because transitioning to a new world is constantly faced with adversity, and loneliness is unavoidable for many newcomers.^49, 51^ Individuals build coping strategies such as constant and unhealthy feeding, shopping, watching TV mindlessly and spending their time anyhow to escape isolation, explains Yanguas et al.^52^ Some students identified calling home as a way of coping in this research. In the general population, the detrimental influence of loneliness on the health of a person has been well studied, and many physical symptoms have been related to stress in the general population, such as suppressed immune system and cardiovascular disease, and psychiatric problems, such as anxiety and depression.^41^

A study reported that loneliness is associated with relatively poor student physical wellbeing and depression.^45^ However, when feeling alone, most participants expressed low self-care, and one suggested a lack of appetite. This was evident in this study as participants expressed self-medication, inadequate self-care, lack of appetite and poor sleep. Some participants expressed a loss of motivation, which impacted their study.

During the interview, participants expressed various coping mechanisms when asked. The main theme included coping mechanisms with sub-themes, religious support, socialisation, and well-being. Some students expressed engaging with religious groups for support in minimising their loneliness. Previous studies on loneliness among university students have examined correlations with the use of attachment, mental illness, and academic success of the social media internet and smartphone.^3, 53^ Most participants identified calling their families daily to keep abreast of what is happening at home, although this could cause more anxiety.

In this research, most students expressed challenges in UK education, such as IT skills. The participant emphasised universities teaching basic skills and supporting students whose learning styles are different in the country. Loneliness can influence the student’s low esteem and interpersonal relationships, explains Mushtaq et al.^54^ It is known that support for guidance can be a valuable instrument to motivate students to engage in class.^51^ Lin and Kingminghae^50^ state that international students troubled by loneliness need help and resources.

Psychological wellbeing is a concept used in positive psychology, synonymous with a positive mood, pleasure and happiness in one’s life.^41^ The theme found in this study was perceived psychological factors, and subthemes were withdrawal, distraction, and detachment. The participants felt detached and isolated from family, making their loneliness worse. Loneliness and depression^7, 8^ tend to be predominant in people with elevated attachment distress over separation, according to a literature review.^55^ Ainsworth and Bowlby^56^ and Ainsworth et al.^57^ work explain that attachment is affection and a bond relating to another person.^58–60^ They believed that children’s initial commitment to their caregivers significantly influences their lives. The attachment of an infant to his caregiver or mother is helpful for his or her life. Again, this hypothesis promoted the belief that being alone could cause difficulties for people. He defined the need to belong. ^41^

Some factors highlighted in this theme were food, weather, navigation, homesickness, support and language barrier. Values and norms are represented in our minds and help with adjustments. On the contrary, positive adjustment to separation from home can enhance the experience, nurture new and existing close relationships, promote resiliency, and prepare students for future success.^61^ According to Ng, Wang and Chan,^62^ evidence indicates the influence of social care on psychological adaptation functions through sociocultural adaptation. This is evidenced in the data collected, making the participants compare the lifestyle and culture of their original country with the host country (UK) in the context of loneliness.^20^ International students must become biculturally competent and adhere to culture, considering their Nigerian cultural identities and values.^63^ (Bii, 2015).

In a culture other than their own, students entering universities must contend with new social and educational institutions, attitudes and aspirations, and the difficulties of adaptation common to students in general.^17^ Most participants in this research expressed homesickness as a contributing lead to loneliness. Moving to another part of the planet is stressful; away from family and friends, from a common language, culture and way of life. ^21^ They are sometimes referred to as cultural shock.^64^ Combined with affordability, convenience will give off-campus students a more significant opportunity to select from the neighbourhood’s accessible food^65^ sources. On the other hand, other reports have found that students living off-campus are more at risk than those living on campus.^66^

## Limitation to the study

This research was about loneliness, and although the researchers chose the topic before the global pandemic COVID-19, it saw that COVID-19 played a significant role in projecting loneliness severity from the participants. This has a limiting effect on the number of participants for the data collection. Interviews were performed online in adherence to COVID-19, which caused IT glitches and interview cancellations due to other factors. A total of 8 participants of Nigerian origin were interviewed for this research which is a relatively small number compared to the total number of international students studying at the institution where the study was conducted. The participants interviewed were of Yoruba, Benin and Igbo origin; therefore, the results cannot represent the entire Nigerian population because it is missing the Hausa tribe. The danger in using a small participant group is that this does not represent the diversity of the student population.

## Future Research

There is a significant need for future research to explore international students from various countries other than Nigerian origin.

## Conclusion and Recommendation

The researcher’s findings highlighted the impact of loneliness on Nigerian students. This indicated that loneliness is an important topic. The findings provided essential information about Nigerian students coming to study abroad. It means that efforts to help cope with loneliness must take into consideration more emphasis on understanding each student’s cultural background.

Feelings of loneliness can be the outcome of emotional, psychological, physical, or mental health. Therefore, there should be a need to support the students going through these factors. Assisting international students to examine students’ bicultural realities to understand how clients internalise and adhere to culture in light of their Nigerian cultural identities and values.^63^

Students expressed the problematic aspect of adaptation to the UK weather as this greatly impacted their studies. The time difference in winter was also seen as challenging to some participants as they informed that the Nigerian weather is mostly summer. Seemingly, the structure and consistency of student life were not parallel to their expectations in the UK. This transition was challenging for the Nigerian students as they moved away from a country where everyone is ‘your family’ and dependent on each other.

After a critical review of various literature and the outcome of the interviews evidenced by the above findings, it is concluded that loneliness had impacted Nigerian students studying at a university in London.

### Recommendations

The following recommendations are designed to enhance the support system for Nigerian international students, addressing their academic, social, and psychological needs.

### The University

#### Comprehensive Support Services

Ensure that students receive mental health, social, and psychological support from the time of enrolment. This includes accessible mental health services tailored to the needs of international students, with culturally sensitive support for coping with loneliness and other issues.

#### Academic Assistance

Identify students’ learning needs and offer support in IT, referencing, Turnitin submissions, and language support programs to help with communication barriers and prevent plagiarism.

#### Employment and Career Guidance

Provide information on part-time job opportunities, offer career advice early in the course, and conduct regular meetings to update students on employability and application processes.

#### Affordable Living Resources

Work with local agencies to identify affordable restaurants and food shops, helping students manage living costs.

#### Enhanced Orientation and Induction

Offer comprehensive and detailed orientation programs that cover academic, social, and cultural adaptation aspects to better prepare students for the challenges they might face.

#### Support Networks and Mentorship

Facilitate the creation of support networks and peer groups specifically for international students, including mentorship programs where new students are paired with senior students for guidance.

#### Cultural Sensitivity and Community Building

Provide cultural sensitivity training for staff and faculty to create an inclusive environment. Organise regular social events, cultural exchange programs, and group activities to build a sense of community and belonging.

#### Communication and Integration Platforms

Create platforms for students to communicate with both home and international peers, and organise social evenings to bring together diverse cultures.

### The Students

#### Proactive Support Seeking

Seek support upon arrival in the UK to navigate university resources and requirements.

#### Emotional and Cultural Preparation

Prepare emotionally for the transition, conduct background research on UK culture, enhance language skills, and become informed before arriving.

#### Social Group Formation and Integration

Create social groups, such as WhatsApp, to connect with peers from similar backgrounds and engage with the host country’s culture.

Implementing these recommendations will enable the universities to create a more supportive and inclusive environment, helping Nigerian international students integrate, thrive academically, and maintain their well-being.

## Data Availability

All data are already provided as part of the submitted article

